# Contextualizing Women’s Birth Experiences at Hospitals in Conflict-Affected Settings in Northeast Nigeria and Eastern Democratic Republic of Congo: A Mixed Methods Comparative Case Study

**DOI:** 10.64898/2026.07.21.26358548

**Authors:** Sussan Israel-Isah, Meighan Mary, Rosine Bigirinama, Salomine Ekambi, Christian Chiribagula, Charity Pring’ar Maina, Kazeem Olalekan Ayodeji, Rejoice Helma Abimiku, Shatha Elnakib, Pacifique Mwene-Batu, Ghislain Bisimwa, Rifkatu Sunday Aimu, Gaylord Amani, George Odonye, Christine Chimanuka, Emilia Ngozi Iwu, Hannah Tappis

## Abstract

**Introduction:** In conflict-affected settings, insecurity, poor health systems, and displacement hinder women’s access to timely, quality maternal and newborn care. In Northeastern Nigeria and Eastern DRC, access to quality intrapartum care remains limited, contributing to preventable deaths. This study compares the quality of intrapartum care in rural hospitals in Yobe State (Nigeria) and North and South Kivu (DRC), using, facility assessments, provider perspectives and women’s experiences to inform women-centered improvements in maternal health.

**Methods:** A comparative mixed-methods case study was conducted across three rural public hospitals selected from the most insecure districts in Yobe State and North and South Kivu. Data were collected through facility assessments, semi-structured interviews with providers, and focus group discussions with postpartum women and triangulated for an in-depth understanding of care quality in these settings.

**Results:** Women’s childbirth experiences across the three conflict-affected settings were shaped by a combination of under-resourced but functioning facilities, financial barriers, sociocultural norms, and interactions with health personnel. While structural limitations such as overcrowding, limited privacy and infrastructures, and cost barriers were reported in all settings, the degree of support from external actors and local health governance shaped key differences in experience. Autonomy in birth decisions was often limited, and perceptions of communication, respect, and emotional support varied. Nonetheless, across all contexts, compassionate provider behavior was consistently valued by women and contributed to more positive experiences, even in strained environments.

**Conclusions:** Improving maternal care in these settings requires systemic reforms that combine infrastructure and workforce investment with attention to sociocultural norms, women’s autonomy, and equity.

## Introduction

The confluence of insecurity weakened health systems, and displacement in conflict-affected settings have important implications for women’s access to timely and quality maternal and newborn services. Barriers such as long distances, (1–3), insecurity, resource scarcity, and shortages of skilled health personnel hinder the delivery of timely, safe, and respectful maternal and newborn health (MNH) services, especially in rural areas (3–5). These fragile and often substandard conditions surrounding childbirth in conflict-affected low- and middle-income countries (LMICs) represent a pressing global concern.

In Northeastern Nigeria and Eastern Democratic Republic of Congo (DRC), similar barriers have resulted in limited access to high-quality intrapartum care, perpetuating preventable deaths and undermining efforts to improve maternal and newborn survival (3). Fear, trauma, and social disruption brought about by prolonged violence and instability also influence maternity care (6,7). Zhang et al. found that in the DRC, self-reported insecurity among mothers was associated with changes in maternal health-seeking behavior, notably delays in accessing care and a preference for home births due to fear (6). In Yobe State, Nigeria, Ager et al. (2015) reported that the Boko Haram insurgency severely disrupted health system functionality, reducing the availability and reach of services for women and children (8).

Beyond these structural obstacles, socio-cultural norms strongly influence the way women experience childbirth in these contexts. In DRC, Nigeria, and across sub-Saharan African countries, traditional beliefs and gender norms influence decisions about where and how to give birth (9–12). Fear of a medicalized birth - particularly a caesarean section - combined with the expectation of family support or preference for informal care based on traditional practices, can discourage institutional care (13). In patriarchal environments, women’s autonomy in childbirth decisions is often limited, with husbands, elders or community figures playing a central role in determining care-seeking behavior (14–16). Cultural perceptions of pain, modesty and the role of care providers also influence how women interpret and respond to hospital birth experiences (13).

While much attention has been paid to maternal mortality trends, service delivery challenges, and access to care in conflict-affected areas, less is known about the lived experiences and perspectives of women giving birth in these fragile systems(17). There is a critical gap in understanding how women perceive the care they receive, what cultural or systemic barriers they face, and how conflict influences their decision-making, autonomy, and trust in the health system. Understanding what they value, fear, endure, and expect are essential for improving the quality of care and designing health system changes which are responsive to the realities of women’s lives in crisis contexts.

This study addressed these gaps by comparing the quality of intrapartum care in three rural hospitals across conflict-affected regions of Northeastern Nigeria and Eastern DRC. Capturing women’s experiences of maternal health care, alongside provider perspectives and facility assessments, contextualizes and informs women-centered interventions poised to improve maternal health outcomes in conflict-affected contexts.

## Methods

A comparative mixed methods case study was conducted to understand women’s experience of intrapartum care as part of a larger research portfolio assessing the quality of intrapartum care provided at health facilities in conflict-affected settings of Northeastern Nigeria and Eastern DRC. A case study approach was employed to enable an in-depth interrogation of the complex and rich experiences of birth (18). By triangulating different methods and perspectives, we were able to contextualize the lived realities of women delivering in conflict-affected contexts. Each case was defined as a rural public hospital within the most conflict-affected settings in 1) Yobe State, Nigeria, 2) North Kivu, DRC, and 3) South Kivu, DRC. Standardization of study questionnaires allowed comparisons between the three cases. Health facility assessments, health provider interviews, and focus group discussions (FGDs) with patients were conducted from February-October 2023. The study protocol was reviewed and approved by the Johns Hopkins Bloomberg School of Public Health Institutional Review Board (IRB No:21922), Yobe State Health Research Ethics Commission in Nigeria (MOH/GEN/747/VOL.1), and Université Catholique de Bukavu Ethical Committee in the DRC (UCB/CIES/NC/011/2023). All data were collected, processed and analyzed in strict accordance with ethical standards and regulations.

### Study Settings

Our study was conducted in Yobe State, Nigeria; and North Kivu and South Kivu, DRC (Figure 1). Within each region, we purposively selected one of the most conflict-affected districts (local government areas in Nigeria; health zones in the DRC) in collaboration with local Ministry of Health stakeholders. Only one hospital served each selected district and was selected as our study site. District and hospital names are de-identified hereafter to preserve confidentiality.

**Fig. 1.**
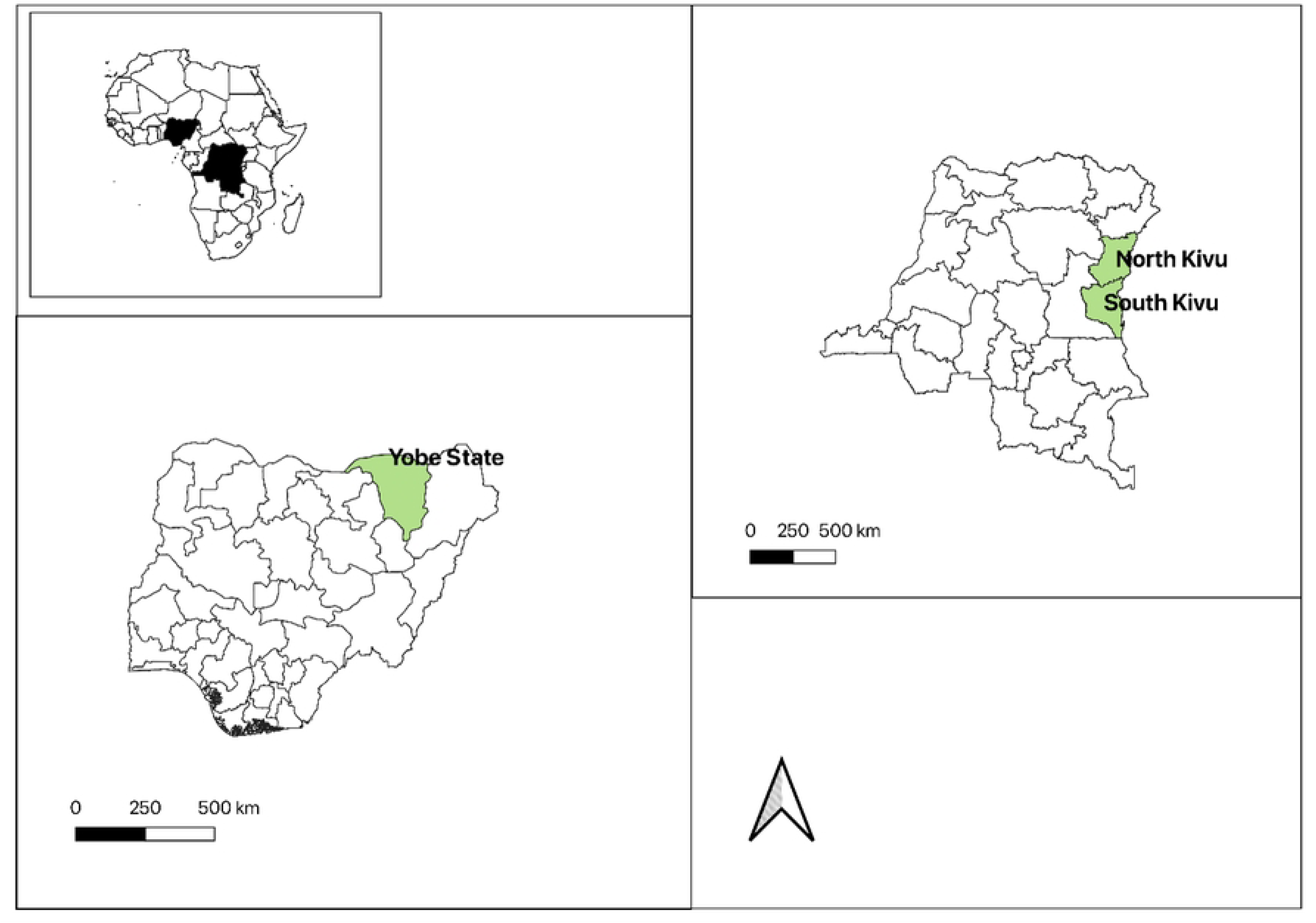
Study sites.

The selected district in Yobe State, Nigeria (Case 1) is one of the areas most affected by the Boko Haram insurgency (19), located approximately 55 kilometers from Damaturu, the Yobe State capital. The routes connecting the district to Damaturu have been subject to sporadic attacks and ambushes, hindering humanitarian access and supply chains (20). Yobe State is home to approximately 2,075,257 returnees (who were previously displaced due to armed conflict), a significant portion reside within the selected district in overcrowded settlements with limited access to food, shelter, clean water, and essential services (21,22). Health infrastructure in the selected district in Yobe State has been severely impacted by the conflict, with many facilities damaged, destroyed, or abandoned due to insecurity (8). Health personnel are often targeted by insurgents; they face significant challenges working in this unstable environment with limited resources and inadequate security. Health facilities are understaffed, lack essential medical supplies, and have a limited number of facilities to provide comprehensive maternal and newborn care (7).

North Kivu (Case 2) and South Kivu (Case 3) are two of the provinces most affected by conflict in the eastern DRC. In North Kivu, the humanitarian situation is critical; in 2023, North Kivu was home to over 2.5 million internally displaced people, many of whom live in precarious conditions with limited access to basic health services (23). Health infrastructure is regularly affected by active conflict-related destruction or abandonment. Healthcare personnel often work in unstable environments with limited resources. In the rural districts of North Kivu, health facilities are generally understaffed and under-equipped, and many lack the capacity to provide emergency obstetric and newborn care. The selected district (health zone) is located over 100 km from the provincial capital, Goma, and is home to a considerable number of internally displaced persons (IDPs) and refugees.

South Kivu, although similarly affected by the conflict, faces slightly different dynamics marked by localized insecurity, structural poverty and weak governance. Armed groups remain active, particularly in rural territories where health infrastructure is scarce, and health districts cover large, hard-to-reach populations. South Kivu is also home to over 1.5 million displaced people regularly affected by recurrent epidemics, sexual violence, and food insecurity (23). In rural hospitals, access to skilled birth attendants and life-saving interventions such as caesarean section and blood transfusion is often limited. The selected *district* is geographically difficult to access due to insecurity and impassable roads.

### Sample

Within each case, one health facility assessment was conducted along with structured interviews with up to eight maternal health providers and two FGDs with women who recently gave birth at the facility. Sample size calculations for health provider interviews were calculated to assess differences in perspectives by health facility for the larger research portfolio assessing quality of care in high volume health facilities across Yobe State, Nigeria and North- and South-Kivu, DRC; our descriptive comparative study included data from all completed interviews in the selected sites. Given the homogeneity of the populations served by each hospital, we estimated that the perspectives from 6-10 people invited in each FGD would be sufficient to reach saturation (24,25); a second FGD was added to ensure we captured any difference or nuance among women who experienced complications.

Health providers were eligible to participate if they were responsible for antenatal, intrapartum, and/or postpartum services. Health workers with no direct involvement in maternal health service delivery (i.e., serving only administrative roles or working in other departments) were excluded from the study. If more than eight health providers were eligible at the time of data collection, participants were selected at random.

Two FGDs were also conducted at each facility: 1.), with women who experienced uncomplicated delivery and 2.) with women who experienced complications during childbirth to ensure representation from the breadth of experiences. Women were eligible to participate if they were aged 18 years or older and delivered a live birth at the health facility within three months prior to data collection. Women with immediate or extended family members working as maternity care providers at the health facility were excluded from the study. A random sample of up to 10 women were selected to participate in each FGD. Table 1 outlines the participant characteristics for each case.

### Data Collection

Data collection was conducted by teams of trained data collectors: 18 in Yobe State, 22 in South Kivu, and 18 in North Kivu. Gender balance was ensured across all three sites, with a gender ratio of 1:1. Data collectors were public health professionals or clinicians with at least two years’ research experience. All data collectors completed comprehensive 7-day training on study procedures, including research ethics and data quality assurance.

### Health Facility Assessment

The district-level chief of health and the Medical Director of each facility received a letter from the respective Ministries of Health to obtain approval for data collection. Upon approval, data collection teams requested support from personnel in charge of the maternity ward in completing the health facility assessment. A standardized tool was developed from the Demographic and Health Surveys (DHS) Program Service Provision Assessment (SPA) (26) and NEST360/UNICEF Health Facility Assessment Toolkit (27) to include an inventory of medicines, supplies, and equipment, a review of patient records, and information on staffing, infrastructure, and maternal and neonatal health protocols. Completion of the health facility assessment took approximately two hours.

### Structured Interviews with Health Providers

To facilitate the health provider interviews, the Medical Director and/or the heads of the MNH departments introduced their personnel to the study team who explained the purpose of the study. All eligible maternal and newborn health service providers on duty at the time of data collection were invited to participate. Interviews were scheduled at convenient times to not interfere with patient care duties.

Interviews were held in locations with visual and auditory privacy (e.g., private offices or conference rooms). Written consent was obtained prior to initiating all interviews. Adapted from the DHS Program SPA (26), a structured interview tool was employed to document health provider knowledge, attitudes, practices, and challenges faced in MNH service provision, along with health provider experiences and considerations related to conflict-affected contexts. On average, interviews lasted up to 30 minutes and were conducted in English (Nigeria) or French (DRC).

### Focus Group Discussions with Women Who Recently Gave Birth

FGD participants were recruited using the health facility registers. A list of up to 30 patients who met eligibility criteria (three times the anticipated number of participants for each FGD) was created with their contact information. Study team members randomly selected from the list and health facility staff in Case 1 (Yobe, Nigeria) and community health workers in Cases 2 and 3 (North-Kivu and South-Kivu, DRC) made the first contact via phone to introduce and inform patients of the study. When interest in participating was expressed, the study team fully described the study, verified their eligibility, and invited them to participate in the FGD. Patients were randomly selected and contacted until the desired sample size was reached for each FGD (6-10 participants).

FGDs were held in a neutral private and confidential location to ensure open and frank participation (e.g., school classroom, church, health zone central office meeting room). Upon arrival, written consent was obtained from each participant. FGDs were conducted by teams of two in local languages (Nigeria: Hausa; DRC: Swahili): one facilitator and one notetaker who were both fluent in the local languages. The facilitator established discussion rules with the participants and responded to any remaining questions before initiating the focus group using a piloted semi-structured guide. The FGD guide was adapted from WHO tools used in a multi-country study on how women are treated during facility-based childbirth (28). All FGDs were recorded and lasted approximately 90 minutes in length.

### Analysis

The health facility assessments and health provider interviews were conducted using the REDCap electronic data capture application (29,30) on tablet computers. Data was uploaded to a central secure server for quality assurance checks and cleaning within 24 hours of data collection or as soon as a reliable internet connection could be established. Quantitative data were analyzed using Stata software (version 18) (31). FGD audio recordings were transcribed verbatim and translated into English (Case 1) and French (Case 2 and 3) and verified for accuracy and fidelity by study co-investigators. Verified transcripts were uploaded into Dedoose software (32) for qualitative coding.

Analysis focused on five domains of the WHO framework for the quality of MNH care (33): two cross-cutting domains including essential physical resources and competent and motivated human resources, along with three ‘experience of care’ domains (i.e., effective communication, respect and preservation of dignity, and emotional support). Table 1 summarizes key quality of care constructs that were assessed for each component of the framework.

**Table 1:** Quality of Care Constructs.

|  | Key constructs collected |  |  |
| --- | --- | --- | --- |
|  | Health facility assessment | Health provider interview | Focus group discussion |
| Essential physical resources | Status of health facility infrastructure<br>Availability of medicines, supplies, and equipment | Health provider perspective of work environment | Patient perspective of birth environment |
| Competent and motivated human resources | • Availability of skilled birth attendants | • Training and perceived competencies of health providers | • N/A* |
| Respect and preservation of dignity | N/A | • Provider training on respectful care | • Autonomy in decision-making<br>• Privacy and confidentiality during labor and delivery<br>• Mistreatment during labor and delivery |
| Effective communication | N/A | N/A | • Effective interaction with health personnel |
| Emotional support | • Policies on birth companions | N/A | • Birth companion support<br>• Health provider support |

\* NA: not applicable

Qualitative transcripts were coded by a team of two co-investigators in Nigeria and DRC, respectively. A priori shared qualitative codebook was developed deductively using key constructs related to each quality-of-care domain addressed in the FGD guide. However, an iterative process was employed to refine the codebook and add codes or sub-codes that were derived naturally from the data. Thematic content analysis was employed to synthesize findings by key constructs and domains for each case, with additional disaggregation of the experience of care constructs by delivery type (complicated vs uncomplicated) (34). Descriptive statistics of data from the health facility assessment and health provider interview tools were also performed by case.

Case descriptions were developed detailing key characteristics of each hospital system. In addition, key constructs related to patients’ experience of care were triangulated with the cross-cutting domains (physical and human resources) to contextualize the perceived quality of care among women with complicated and uncomplicated deliveries in each case. Health provider and patient perspectives of the security context were also summarized. Individual case findings were compared providing in-depth insights on the experience of childbirth across three different crisis-affected contexts.

### Positionality

Reflexivity was embedded throughout the study period. Data collectors in each country received training on practicing interpersonal and cultural reflexivity during and after health provider interviews and FGDs, with the aim of minimizing subjectivity and addressing potential power imbalances. All co-authors were actively involved in the analysis and interpretation of the data. The research team comprised members from the DRC, Nigeria, and the United States, each bringing personal and/or professional expertise in maternity care. Regular team meetings served as a forum for collective reflexivity, allowing the team to critically assess coding decisions and emergent themes while addressing possible contextual and personal biases in the interpretation process.

## Results

### 1. Participant characteristics

Table 2 outlines the participant characteristics for each case.

**Table 2.** Participant Characteristics.

|  | Case Study 1: Yobe<br>State, Nigeria | Case Study 2:<br>North Kivu, DRC | Case Study 3:<br>South Kivu, DRC |
| --- | --- | --- | --- |
| *Health provider interviews* | N=8 | N=8 | N=5 |
| Sex: |  |  |  |
| Male | 0 (0%) | 3 (38%) | 2 (40%) |
| Female | 8 (100%) | 5 (63%) | 3 (60%) |
| Health provider cadre: |  |  |  |
| Midwife-Nurse | 8 (100%) | 7 (88%) | 3 (60%) |
| Doctor | 0 (0%) | 1 (13%) | 2 (40%) |
| Held a management position | 3 (38%) | 3 (38%) | 2 (40%) |
| Years working in facility: |  |  |  |
| <2years | 4 (50%) | 2 (25%) | 3 (60%) |
| 2+years | 4 (50%) | 6 (75%) | 2 (40%) |
| **Focus group discussions** | **N=20** | **N=20** | **N=20** |
| Delivery type: |  |  |  |
| Complicated delivery | 10 (50%) | 10 (50%) | 10 (50%) |
| Uncomplicated delivery | 10 (50%) | 10 (50%) | 10 (50%) |
| Median Age (IQR): | 20 (19-21) | 25.5 (21.5 – 29.5) | 20.5 (19.0 – 23.5) |

### 2. Case summaries

Key characteristics of each selected case are described in Table 3. Case 1 in Yobe, Nigeria is a hospital that is managed by the State Ministry of Health (MoH) that serves IDPs and the surrounding host communities. Antenatal care and Comprehensive Emergency Obstetric and Newborn Care (CEmONC) are offered. No fees are charged for routine vaginal deliveries or any medicines required for maternity care. However, patients or their families are expected to purchase needed supplies for delivery. Patients undergoing a cesarean section are expected to pay for services, yet those with lower economic status may be exempt from payment or be offered services at a discounted fee and fee is not required before treatment, especially in cases of obstetric emergencies.

Case 2 in North Kivu, DRC, is a state hospital rehabilitated and supported by an international humanitarian organization. It serves a catchment population of 508,969 including IDPs, refugees, and the host population. Antenatal services are not provided at this facility; however, CEmONC services are available at no cost to patients. Additional supplies and medicines required during labor and delivery are also covered by the humanitarian organization.

Case 3 in South Kivu, DRC is a hospital managed by the Provincial MoH. Serving a catchment population of 187,201, it provides antenatal services and CEmONC to IDPs and the host population. In Case 3, fees are charged for both routine vaginal and cesarean deliveries along with the required medicines for maternity care. However, patients and their families are not required to pay for additional supplies during labor and delivery, nor is payment required before treatment during obstetric emergencies. Internally displaced families are exempt from all maternal health service fees. In addition, in-kind payment is accepted from patients who are unable to pay fees associated with their maternity care.

**Table 3:** Case characteristics.

| Characteristics | Case 1: Yobe, Nigeria | Case 2: North Kivu, DRC | Case 3: South Kivu, DRC |
| --- | --- | --- | --- |
| Hospital managing authority | State MoH | Provincial MoH supported by humanitarian organization | Provincial MoH |
| Catchment population | >198,000<sup>a</sup> | 508,969 | 187,201 |
| Populations served: |  |  |  |
| IDPs from within the province/state | Yes | Yes | Yes |
| IDPs from outside province/state | Yes | Yes | No |
| Refugees | No | Yes | No |
| Host communities | Yes | Yes | Yes |
| **Service Provision** |  |  |  |
| Antenatal care provided | Yes | No | Yes |
| CEmONC capabilities | Yes | Yes | Yes |
| Allows birth companions to be present during labor and delivery | No | No | No |
| **Payment of Services** |  |  |  |
| Charges fees for: |  |  |  |
| Routine vaginal delivery | No | No | Yes |
| Cesarean delivery | Yes | No | Yes |
| Medicines for maternity care | No | No | Yes |
| Populations exempt from paying fees for maternal health services: |  |  |  |
| IDPs | No | N/A | Yes |
| Refugees | No | N/A | No |
| Patients who do not have any means to pay | Yes | N/A | No |
| People with physical or cognitive disabilities | No | N/A | No |
| Procedure if a patient is unable to pay for fees associated with maternal health care: |  |  |  |
| Fee exempted, no payment expected |  |  |  |
| Fee discounted | Yes | N/A | No |
| In-kind | Yes | N/A | No |
|  | No | N/A | Yes |
| Expect patients or their families to purchase supplies for routine vaginal delivery | Yes | No | No |
| Expect patients and their families to purchase supplies prior to treatment for an obstetric emergency | No | No | No |
| Payment required before a patient can receive treatment during an obstetric emergency | No | No | No |
| Abbreviations: CEmONC: Comprehensive Emergency Obstetric and Neonatal Care; IDP: Internally Displaced Persons; N/A: not available |  |  |  |
| <sup>a</sup> Formal catchment population not calculated for the referral hospital; we report the local government area (LGA) population for comparison, noting that the hospital often serves patients residing outside its LGA. |  |  |  |

While these services and policies were reported at the hospital, FGD participants shared conflicting accounts reflecting how these policies were translated into practice. Financial constraints related to each hospital’s payment policies were a major obstacle during their hospital deliveries. For example, maternity services were reported to be free of charge in North Kivu, whereas is South Kivu (Case 3) women reported being unable to leave the hospital until their bills were settled. Additionally, in Case 3 (South Kivu), participants added that they felt pressured to have cesarean deliveries, suspecting that the frequent recommendations were financially motivated rather than clinically necessary. Such perceptions created suspicion and reduced confidence and satisfaction in facility-based care. Similarly, in Yobe (Case 1), participants shared that people in their communities preferred avoiding hospital deliveries due to prior experience with financial obligations for intrapartum care. In fact, the majority of participants that experienced complications during delivery only attended the hospital following a referral and/or due to the onset of danger signs.

> *“My neighbor…doesn’t allow his wives to come to the hospital; he will say they want to make him bankrupt. He said his sisters and mother usually give birth at home, why won’t his wives? Even if they come, he will be insulting and fighting the health workers.” – Participant who had an uncomplicated delivery in Yobe, Nigeria (Case 1)*
>
> *“I was billed even though I went straight to the delivery room without going through the labor room. I didn’t receive any post-delivery medication. I asked them to reduce the bill, but they refused.” - Participant who had an uncomplicated delivery in South Kivu, DRC (Case 3)*

### 3. Cross-cutting quality of care domains: Physical and human resources

Each case’s available physical resources (Table 4) were also triangulated with FGD participants’ reports of their birthing environments. The two cases in DRC (Case 2 and 3) had one labor room and one delivery room available and the case in Yobe, Nigeria (Case 1) had two delivery rooms. The condition of delivery rooms varied across cases. In Yobe and South Kivu, each delivery room was private with one delivery bed. However, in North Kivu, the delivery room was shared, with curtains to maintain visual privacy between four delivery beds.

North Kivu FGD participants described the delivery rooms as overcrowded with limited space and insufficient number of beds, at times resulting in multiple women sharing beds. The Yobe and South Kivu FGDs, participants also mentioned limited availability of labor and delivery beds in the hospital. Despite reported labor and delivery ward crowdedness, the delivery environments in all cases were generally described as clean and organized, and the staff were commended for maintaining good hygiene.

In South Kivu, FGD participants reported a shortage of painkillers” (analgesics). One participant shared:

> *“I’ve said it all, but the problem with this hospital is the lack of medication and the high bill, we’re very poor in our community… We are only entitled to one tablet of paracetamol and one Ibuprofen for a week. Whether you are healed or not, they don’t care - Participant who had a complicated delivery in South Kivu (Case 3)*

While the health facility assessment did not include analgesics, each case reported the availability of essential maternal health medicines (Table 4).

The experience of FGD participants was also validated by health providers interviewed in each case. When asked about their work environment, health providers across all cases most frequently mentioned limited health facility infrastructures (e.g., limited space and number of delivery beds) and high workload as ongoing challenges in providing high quality maternal health care. Health providers in Yobe and South Kivu cases also discussed issues with the lack of and/or stockout of medicines or equipment, while North Kivu health providers highlighted patient confidentiality during delivery as a primary concern.

The staffing and availability of qualified maternal health personnel were also assessed. Yobe and North Kivu cases both had over 30 nurse-midwives on staff with 5 and 2 doctors at each maternity unit, respectively. Staffing for the South Kivu was much smaller, with only 3 doctors and 3 midwives

However, regardless of total staff size, only 1–2 maternal health providers were scheduled per shift (day or night) in Cases 1 and 3. North Kivu (Case 2) stood out, with 12 staff on duty during the day and 11 at night. In Yobe (Case 1), only day shifts have maximum staffing, the night shifts engage only 2 midwives and doctors on call.

Health provider interviews examined training and capacity in maternity care (Table 4). At most, 50% of health providers interviewed in Yobe (Case 1) reported ever receiving an in-service or refresher training on intrapartum care, compared to 37.5% and 40% of health providers in North and South Kivu (Cases 2 and 3), respectively. In addition, very few health providers across all settings reported receiving training on respectful maternity care. Across the three cases, health providers in Yobe (Case 1) felt the most prepared and trained for their current position (88%). In contrast, all health providers interviewed in South Kivu (Case 3) reported performing clinical maternal health tasks they were not previously trained on compared to 50% of health providers in the Yobe and North Kivu cases.

**Table 4:**
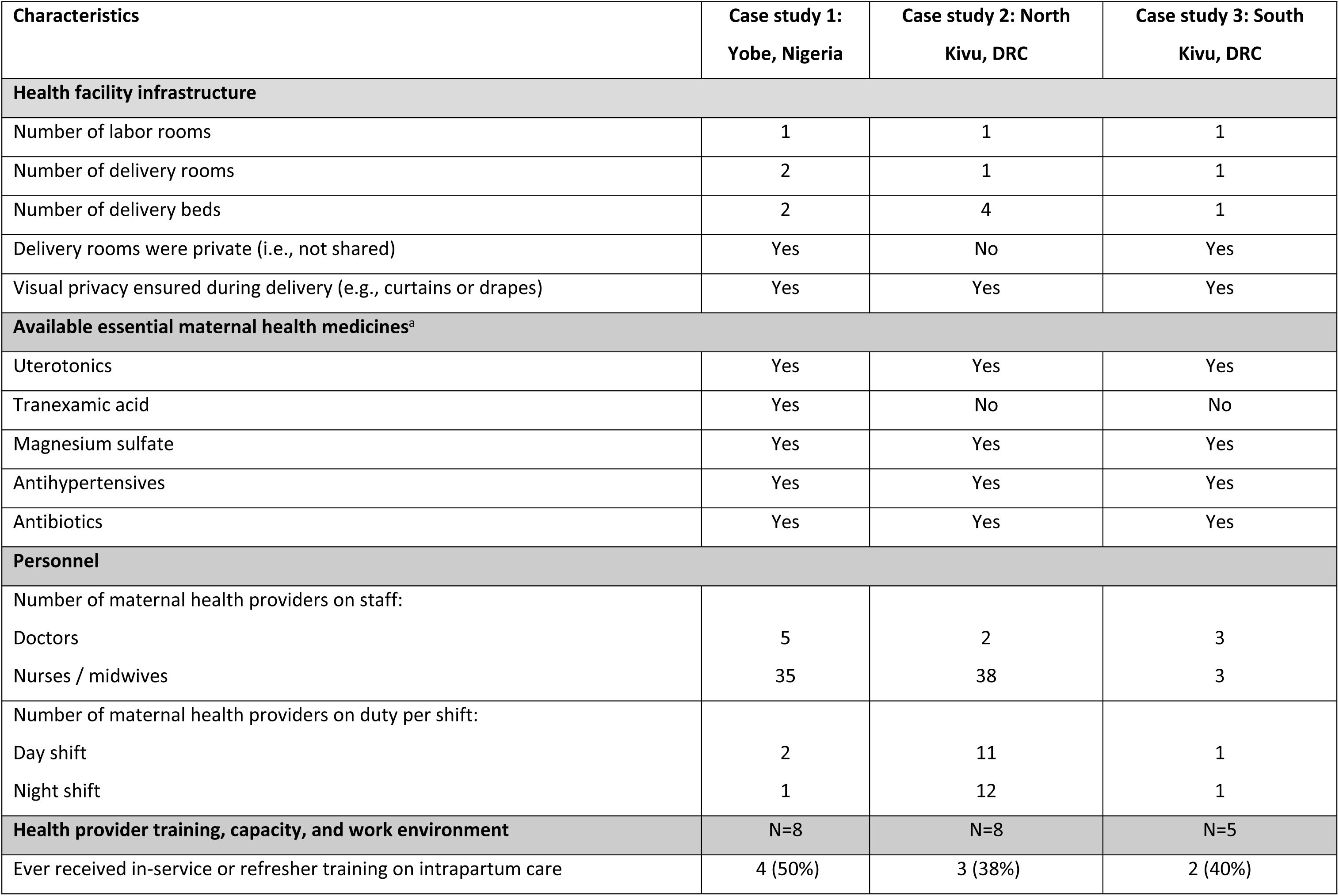
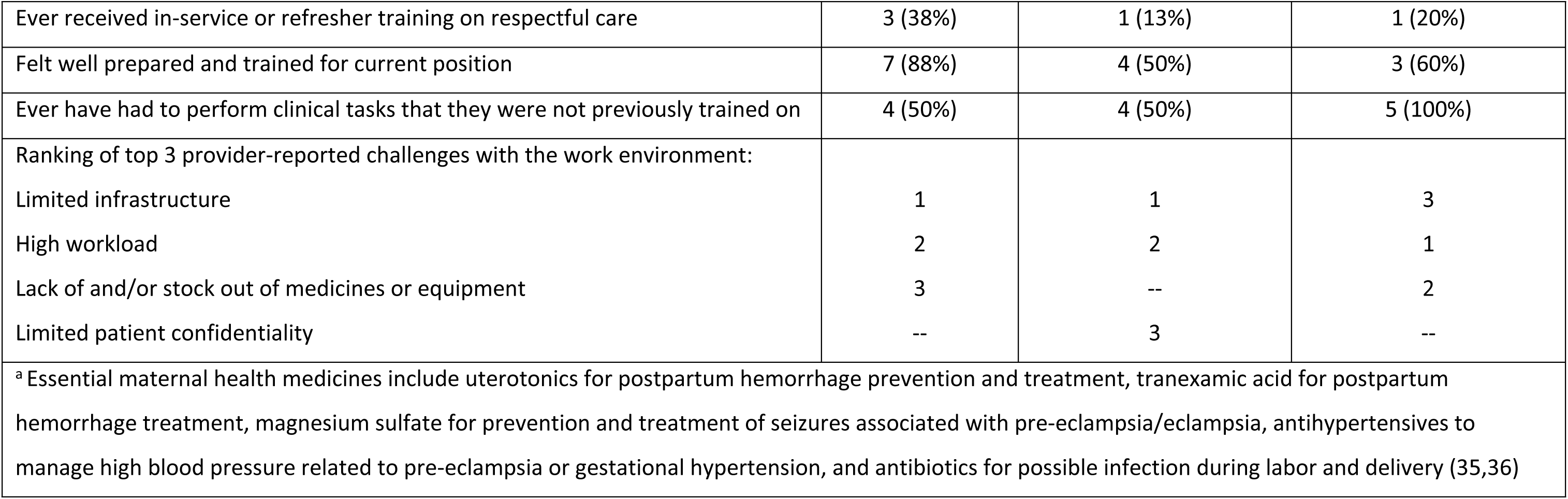
Physical and human resources available in each case.

### 3. Experience of Care Domains

Noting the contextual variations and availability of physical and human resources across the three case studies, we examined women’s experiences of care across three key domains as articulated by women who recently had a complicated or uncomplicated delivery: 1. Respect and preservation of dignity, 2. Effective communication, and 3. Emotional support.

#### Respect and Preservation of Dignity

The extent to which respect and dignity were preserved during labor and delivery was assessed in terms of autonomy in decision-making, the right to privacy, and the experience of mistreatment. Across all cases, FGD participants shared that socio-cultural norms strongly shaped women’s autonomy and decision-making around maternal health care. For instance, in Yobe (Case 1), family pressure and threats from husbands, strongly influenced birth decisions, as men often preferred that their wives follow traditional birthing practices. Some Yobe participants also shared that their husbands only permitted hospital care after intervention from religious or community leaders.

> *“I brought a woman that was sick, she said she feels her baby has died inside her tummy because she doesn’t feel any movement and her husband has warned her not to go to the hospital, if not she will lose her marriage” - Participant who had an uncomplicated delivery in Yobe (Case 1)*

In contrast, FGD participants in South Kivu made delivery decisions independently, typically guided by personal experience or health care advice. While women typically informed their husbands and acknowledged their role as heads of household, they appeared to retain their autonomy. However, when pregnancy complications arose, decision-making was more often shared between women and their husbands.

> *“We decide together whether I should go to the health center in Bukavu [the capital city], or the hospital.” - Participant who had a complicated delivery in South Kivu (Case 3)*

Privacy concerns were mixed, given the difference in delivery room set-up across the 3 cases. Yobe FGD participants mentioned being adequately covered in private labor and delivery areas with restricted access. The South Kivu case also had private delivery rooms; however, FGD participants reported frequent interruptions from trainees and other health personnel passing through the delivery room, which compromised their sense of privacy during childbirth. In the North Kivu case, where the delivery room was shared, participants generally did not express significant concerns about privacy, noting that the curtains in place to offer discretion were sufficient.

Across all cases, participants reported instances of mistreatment. In both North and South Kivu, the nature and degree of verbal and physical mistreatment varied among participants. In cases of complicated deliveries in North Kivu, participants generally described health personnel as respectful and nonviolent. However, some participants who experienced uncomplicated births recounted being insulted or physically mistreated, such as being slapped for not pushing when instructed or for failing to follow provider directions. Similarly, women from Yobe who experienced uncomplicated deliveries mentioned that some health personnel perceive physical reinforcement, such as slapping a laboring woman’s thigh, as a corrective measure to ensure safety during birth rather than an act of mistreatment. Although a few participants disapproved of verbal and physical abuse, across all cases, such practices were justified during critical labor and delivery moments as measures to ensure maternal cooperation to protect the baby.

> *“Sometimes the midwife will slap her on the thigh so they will not kill their child…” - Participant who had an uncomplicated delivery in Yobe (Case1)*
>
> *“When I arrived here, I wasn’t welcomed, they were about to insult me, they asked me how an adult could give birth on the road, I explained the situation to them, but the person taking care of me treated me badly, and I wasn’t happy about it.”- Participant who had an uncomplicated delivery in North Kivu (Case 2)*
>
> *“He has the right to hit you… he’s trying to save your life and the life of your child.”- Participant who had a complicated delivery in South Kivu (Case 3)*

Desired movement during labor and preferred delivery positions were also not respected across settings as delivery positions were chosen by health personnel. Yobe FGD participants had slightly more flexibility in early labor, but women’s choices were not respected during later phases. In South Kivu, health personnel were described as playing a dominant role in determining birthing positions and guiding women throughout the birthing process, leaving little room for women’s preferences.

#### Effective Communication

Participants in each case had varied perceptions of their interaction with health personnel. Across all cases, some women who experienced uncomplicated deliveries reported positive interactions with healthcare personnel. They felt welcomed and appreciated the information provided about their labor progress and when they were fully dilated for delivery. FGD participants mentioned that they were typically provided with explanations for procedures. In Yobe (Case 1), FGD participants accepted the information provided by healthcare staff without question, trusting them as experts and preferring not to challenge their authority by asking follow-up questions.

For women who experienced complicated deliveries, patient-provider interaction became even more crucial. Yobe participants largely described health providers as attentive and communicative, offering reassurance and clarity throughout the process. However, participants in both North and South Kivu felt constrained in their ability to question or challenge healthcare personnel. They perceived compliance as essential to receiving proper care and avoiding potential conflict.

> *“You ask if the baby is doing well, and they tell you that you’re at 2 or 3 cm. They tell you and encourage you.” - - Participant who had complicated delivery in North Kivu (Case 2)*
>
> *“Yes, they did, every test they carried out they told me why”. - Participant who had a complicated delivery in Yobe (Case 1)*

#### Emotional Support

Emotional support during labor and childbirth was determined by the facility’s policies and health personnel attitudes. According to facility policies, none of the cases allowed birth companions to be present during labor and delivery. In all cases, most FGD participants’ experiences reflected these protocols, having given birth without the presence of a family companion.

In North Kivu (Case 2), access to delivery rooms was restricted to only health personnel to prevent outsiders from bringing in unapproved herbal or traditional medicines that may interfere with clinical care standards.

> *“Some people enter with traditional and mystical medicines. This is why access is forbidden. Once you’ve given birth, they’ll just tell the person who accompanied you.” - Participant who had a complicated delivery in North Kivu (Case 2)*

In Yobe (Case 1), lenience in enforcing the birth companion policy was shared, particularly in the case of uncomplicated deliveries. Some FGD participants shared that their mothers or sisters were present in the early stages of labor, usually under specific conditions (e.g., at night or when the delivery ward was not busy). Family members provided outside support, for example, by providing needed supplies, and were only allowed to visit the woman once her condition stabilized.

> *“When they said I will have to do a CS [caesarean section], I asked them to call my mother. When she came, I requested that she stays with me, and they allowed her.” - Participant who had a complicated delivery in Yobe (Case 1)*

Provision of emotional support by health personnel, or lack thereof, also emerged from the discussions. South Kivu FGD participants highlighted the attentiveness of maternity providers in responding to their needs and emotional support during labor and delivery. Similarly, in the North Kivu case, participants emphasized supportive postnatal care, recounting respectful hands-on support during recovery, including help with washing, dressing, and early newborn care. Nonetheless, across both cases, instances were also mentioned where some of the health workers prioritized other patients, leaving women in active labor feeling overlooked. In North Kivu, some FGD participants that had uncomplicated deliveries described negligence from the health providers. Several participants noted that their care was often delayed or neglected, often due to health providers’ favoritism to treat patients that they knew personally and/or the lack of formal referral letters.

In Yobe, experiences with patient-provider interactions were mixed particularly amongst those with complications. Some women felt they were treated kindly and fairly, no matter where they came from, while others felt ignored, tying the neglect to health personnel stress, exhaustion, or perceived judgment of patients.

> *“When I came, the doctor and the in-charge were around, but since my delivery was delayed, they left. So, when it was time to give birth, I called the midwife to come and help me, and she said I should be used to the birthing process by now since I have had 7 children.” - Participant who had a complicated delivery in Yobe (Case 1)*
>
> *“After your delivery, you rest. You wash. The nurses help you. There are those who help you dress. You leave the place feeling very well to go and look after your child.”- Participant who had a complicated delivery in North Kivu (Case 2)*

### 4. Effect of surrounding insecurity

Health provider and patient perspectives on the security situation in each case were also explored. Health provider perception of safety varied widely across settings (Figure 2). Most health providers interviewed at the North Kivu case felt safe from crime and violence at the facility (87.5%) and when traveling to and from the facility (63%). However, safety perception was lower among health providers in the Yobe and South Kivu cases, especially while traveling to and from work (Case 1: 25%, Case 3: 40%).

Health providers also shared insights on the crises’ impact on maternal health service delivery. Health providers in the Yobe case described the destruction of health facilities, the inability to provide consistent health services due to facility closures, and the ongoing psychological pressure on health providers expected to continue work as normal amidst the ongoing insecurity. Health providers in North Kivu shared similar stories and added that mass population displacement during the active conflict has impeded women’s access to maternal health services and left the hospital simultaneously understaffed due to the departure of some health providers. They felt overwhelmed by the influx of people seeking refuge on the hospital premises. Insights from health providers in South Kivu highlight the vulnerability of the hospital which is geographically isolated and hard-to-reach and surrounded by armed groups. They shared challenges with medicine shortages and stockouts and a history of attacks on health providers.

From the patient perspective, security measures implemented in each case played a significant role in FGD participants’ perceptions of safety and protection. In the North and South Kivu cases, the presence of tightly controlled access points, such as mandatory identification checks and restricted entry protocols into the labor ward contributed to a heightened sense of security. Similarly, in the Yobe case, the visibility of security personnel stationed at hospital entrances served as a reassuring presence, reinforcing trust in their safety at the facility.

**Figure 2:**
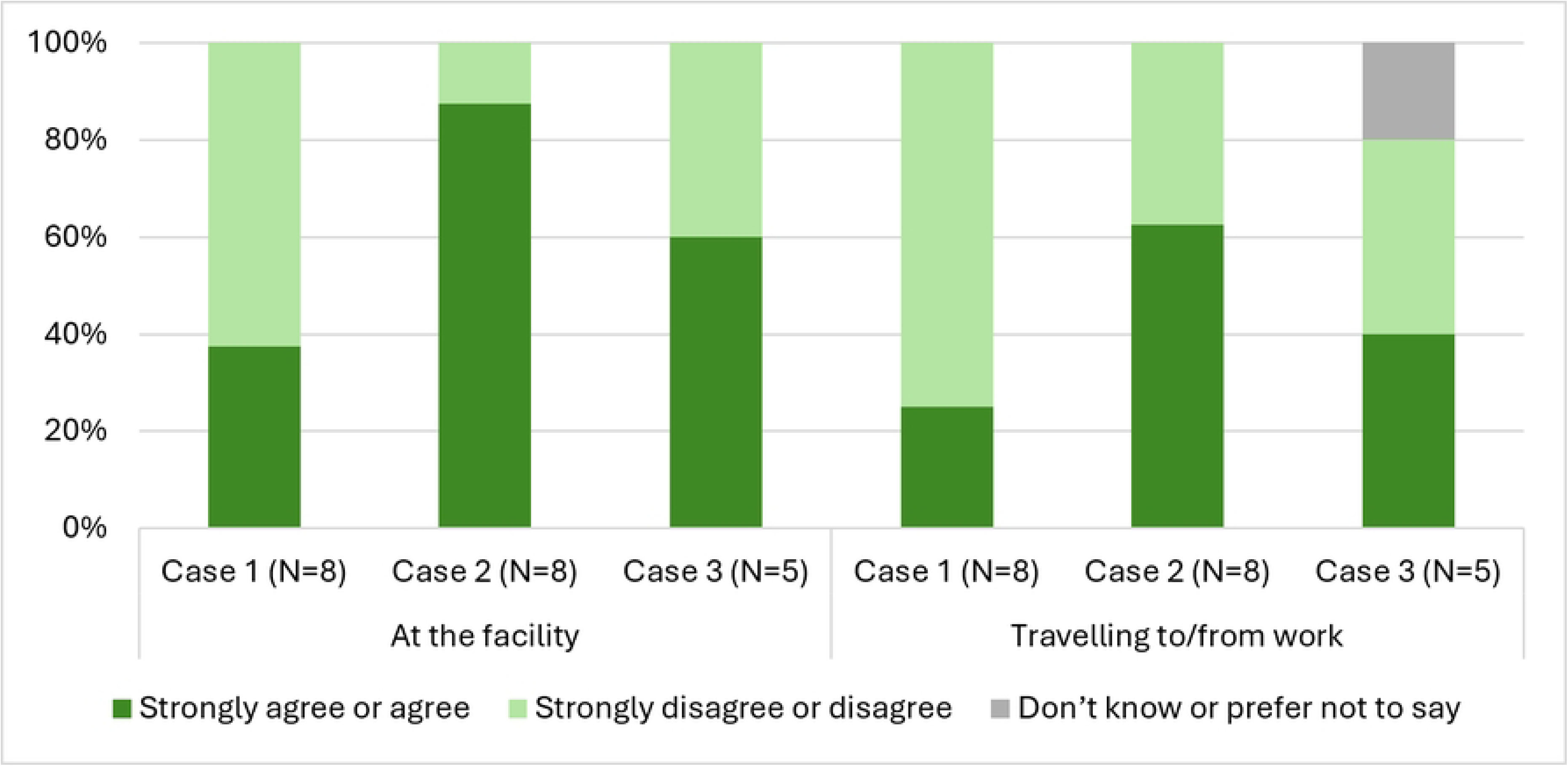
Health Provider’s Perception of Safety While at the Facility or Traveling to/from Work.

## Discussion

This study is the first to triangulate health system, health provider, and patient perspectives to contextualize women’s birth experiences in hospitals situated in crisis-affected areas of Northeast Nigeria and Eastern DRC. Results revealed that participants’ childbirth experiences across all cases were shaped by a combination of factors such as under-resourced yet operational health facilities, sociocultural norms, and interpersonal dynamics with health personnel. For example, while all three cases took place in conflict-affected regions, the differences in the health system context help explain some variation in women’s experiences. Women in Yobe (Case 1) reported relatively better emotional support and communication, potentially linked to the higher proportion of maternity care providers who reported receiving capacity strengthening trainings as well as feeling well prepared to deliver high-quality intrapartum care. Despite an active conflict, relatively better provider–patient relation was found in the North Kivu case which may reflect sustained NGO support that in turn, contributed to improved provider attitudes through accountability mechanisms. In comparison, women in South Kivu faced more pronounced challenges related to privacy and confidentiality, which may in part be due to the hospital’s extreme financial constraints and limited number of maternity providers trained in respectful care. Nonetheless, despite systemic challenges, women in all cases valued compassionate providers who supported their delivery experience, highlighting the importance of both structural improvements and human-centered care to strengthen maternal health in conflict-affected settings.

Effective communication between healthcare providers and pregnant women, a key domain of experience of care, is essential for quality maternal health care (37). While our study found that some women in each case experienced positive, communicative, and attentive interactions during labor and delivery, many highlighted persistent power imbalances that made them reluctant to ask questions and openly dialogue due to fear of compromising their care. This dynamic reflects broader patterns observed in similar settings. A cross-sectional nurse-patient communication study in Nigeria also found that interactions with health workers were often provider-dominated and limited by staff shortages, high workload, and poor communication skills (38). Likewise, direct observations across five countries in East and Southern Africa revealed that providers rarely encouraged women to ask questions, and many women were not informed about procedures (39). Our findings highlight that effective communication is possible in conflict settings yet established social hierarchies and gendered power imbalances in Nigeria and DRC may have influenced provider-patient interactions during labor and delivery.

Our findings related to the respect and preservation of dignity domain, specifically in its women’s decision autonomy component, closely aligned with studies conducted in other conflict-affected regions of sub-Saharan Africa. Like our study, previous research in South Sudan, the DRC, and Somalia have documented how socio-cultural and patriarchal norms influence women’s autonomy in seeking maternal health services (39,40). For instance, in conflict-affected South Sudan, male partners were often reported to have significant control over birth decisions, as they restricted women from delivering in health facilities unless a serious complication arose (41). The normalization of mistreatment, especially during uncomplicated births, has also been widely observed in fragile contexts, as highlighted by our findings. Research in Nigeria and South Sudan reported that women often accepted verbal abuse during childbirth as part of routine care, particularly when such actions were framed as necessary to save the baby or due to understaffed, and overstressed health workers (42,43). Our study reinforces this trend, as physical abuse was often rationalized as helpful rather than harmful. Health provider training in respectful maternal care that emphasizes empathy, cultural sensitivity, patient-centered communication, and respectful care practices should be prioritized. Further investigation on how best to operationalize respectful maternity care models (44) and community engagement strategies in fragile contexts is warranted.

Women’s assessment of emotional support during labor and delivery in Nigeria and DRC was shaped by their limited interaction with family during delivery and the interdiction of birth companions. Hospital policies prohibiting birth companions have also been reported elsewhere (45–49), with the rationale that companions were an unnecessary encumbrance during labor and delivery. However, in many sub-Saharan African countries, including DRC, cultural perceptions discourage male involvement in their partner’s pregnancy (50). Additionally, some women rely on traditional herbal decoctions believed to ease labor, but which may pose health risks (51). According to some FGD respondents, hospitals banned companions partly due to fears that these herbal preparations could be administered clandestinely during delivery. Similar practices and taboos have been documented across sub-Saharan Africa, including Zimbabwe and Nigeria, where companion presence at birth is often restricted for safety or cultural reasons (51,52).

Moreover, within conflict-affected contexts, insecurity, curfews, and displacement often impede the presence of family or birth companions during labor and delivery, even when facilities would allow it. In Yobe, the permanent threat posed by Boko Haram has led to curfews and restricted night-time movement. Meanwhile, in South Kivu, the geographical inaccessibility of the rural hospital combined with lingering insecurity and near-absent transportation options often render birth companionship and familial support during childbirth infeasible. More research is needed to understand how best to respect women’s rights to birth support in accordance with WHO guidelines (53) while mediating additional challenges that health providers may face in doing so, especially in crowded, understaffed, and hard-to-reach maternity wards. In addition, efforts to raise awareness on the benefits of labor and birth companionship (e.g., shorter duration of labor, decreased cesarean sections, increased satisfaction with childbirth (54–58)) are a crucial step to improving the experience and outcomes of childbirth in conflict affected settings.

Our study expanded upon previous research by contextualizing women’s experience of childbirth with objective assessments of the available physical and human resources at their respective health facilities. We found underlying issues with crowded labor and delivery rooms, limited beds, and privacy across all cases. Overcrowding of delivery rooms in Yobe and North Kivu, and the general lack of space reported across cases, point to systemic issues of underinvestment and the inability to meet the needs of the population, albeit with differing root causes. In Yobe and South Kivu cases, underinvestment in health infrastructure is a driving cause while the congestion reported in North Kivu (Case 2) may stem from the hospital’s NGO-supported free care policy, coupled with the extreme influx of IDPs and refugees. This highlights how sustained external funding remains strategical in improving access to and utilization of integrated health care services in humanitarian settings and signals the need for infrastructural improvements that protect privacy and enhance childbirth experience.

Reported high and unaffordable hospital bills also created significant barriers in South Kivu and in Yobe demonstrating stark differences between hospitals supported by non-governmental organizations (as it is for Case 2) and those reliant upon the public health sector (Cases 1 and 3). Parallels are found in Uganda, Nigeria, and Ethiopia where user fees deter many women from seeking timely maternal care, especially those from poorer households (59,60). Our findings reveal a strong need for health policy reforms to ensure maternal health services are free, including childbirth supplies and essential medicines. External subsidies in North Kivu (Case 2) reduced patient costs, while reliance on user fees in public hospitals in Yobe (Case 1) and South Kivu (Case 3) strained access and trust. Eliminating hidden costs and other out-of-pocket expenses are crucial for increasing access to facility-based deliveries and improving health outcomes.

Bohren et al.’s (2015) systematic review synthesizes key domains of women’s experience of childbirth in LMICs, albeit with limited insights from conflict-affected contexts (61). Our findings build upon the evidence base by comparing three distinct conflict-affected settings and highlighting how distinct manifestations of conflict, whether through direct violence, prolonged insecurity, or structural neglect, interacted with local health system capacities and sociocultural dynamics to shape women’s experiences of childbirth (Box 1). Our study identifies ample opportunities for improving maternal health care. Nevertheless, tackling issues such as interpersonal communication, entrenched social and gender biases, and the erosion of trust in health systems to achieve high quality maternity care will require multi-faceted and context-sensitive strategies. In conflict-affected contexts such as the DRC and Nigeria, violence and instability permeate all facets of society, including the provision and experience of maternity care. Our research calls on global and national stakeholders to prioritize more than the direct effects of conflict (e.g., health system functionality, health provider, and patient safety). We urge decision-makers to address the less visible, yet profound, indirect impacts including psychological trauma, chronic stress, and toll of ongoing threats of violence on health providers, women, and their families. These considerations are essential for developing long-term strategies to improve maternity care that not only strengthen systems but humanize care and foster resilience.

Box 1. Contextualizing women’s childbirth experience with conflict-affected contexts

| Case | Summary of triangulated findings |
| --- | --- |
| Case 1: Yobe, Nigeria | The nature of conflict in Yobe was defined by chronic insecurity, curfews, and the persistent threat of Boko Haram insurgency. Women reported limited emotional support, distant provider interactions, and distrust due to financial barriers. While the facility infrastructure remained functional and staff were relatively well-trained, the insecure environment shaped both access to care and overall experience. Curfews and fear limited movement, especially at night, affecting timely access to delivery services and the presence of companions. |
| Case 2: North Kivu, DRC | Women gave birth amid active armed conflict, widespread insecurity, and significant population displacement. Despite the severity of the crisis, NGO support played a critical role in maintaining minimal hospital services, offering free maternal care, and staffing |
|  | facilities. This support may have contributed to better provider–patient interaction and a stronger perception of respectful care. Still, overcrowding in the hospital was common, birth companions were prohibited and familial support during childbirth often absent. The presence of active conflict, paradoxically, did not always go along with the worst experience, highlighting the safeguarding of strong external assistance. |
| Case 3: South Kivu, DRC | The selected health zone faced no active combat, but residual insecurity, vast geographical distances, and infrastructural neglect created major barriers. With no partner support and minimal government funding, the hospital struggled to provide adequate staffing or supplies. Women often delivered without emotional support, in poorly equipped facilities, and bore significant financial burdens. Women’s experience reflected how long-term neglect and isolation, rather than the intensity of conflict itself, can erode care quality and dignity over time. |

Interpretation of our results should be considered while noting several limitations. Our findings are shaped by the perspectives and recall of participants. Given the conflict-affected setting and the fact that some women experienced complicated or traumatic births, recall bias may have influenced their ability to accurately remember and articulate their experiences. Furthermore, FGDs were conducted in local languages and later translated into English or French, leading to possible loss of cultural nuances or subtle meanings.

## Conclusions

This study provides new insights by comparing three distinct conflict-affected contexts and highlighting how conflict, cultural dynamics, and health systems can interact to shape childbirth experiences. We identified how the quality of maternal care in conflict-affected contexts is often shaped by complex and interrelated factors. Women’s experiences of care, including respect, dignity, communication, and emotional support, are influenced not only by individual caregiver behaviors but also by deeply embedded sociocultural norms, limited resources, and systemic neglect, underscoring that respectful maternity care is not simply a matter of individual behavior, but a systemic problem that requires structural change. To improve mothers’ experiences of care in conflict settings, investments must address both structural constraints and sociocultural factors. Strengthening infrastructure, increasing staffing levels, and supporting health care providers must go hand in hand with challenging harmful gender norms and integrating respectful care into policies and training. Without such a multifaceted response, quality maternity care will remain inaccessible to many women in crisis-affected regions.

## Data Availability

Due to the sensitive subject, data is available from the corresponding author on reasonable request and signature of a data sharing agreement.

